# In vivo Characterization of MRI-based T1w/T2w Ratios and Covariance Network in Temporal Lobe Epilepsy

**DOI:** 10.1101/2020.12.03.20243238

**Authors:** Yuchao Jiang, Wei Li, Yingjie Qin, Le Zhang, Xin Tong, Fenglai Xiao, Qiyong Gong, Dong Zhou, Dongmei An, Cheng Luo, Dezhong Yao

## Abstract

Temporal lobe epilepsy (TLE) is the most common type of intractable epilepsy in adults. A novel method based on the ratio of T1-weighted (T1w) and T2-weighted (T2w) magnetic resonance images can investigate brain microstructural changes and how these regional changes interact with each other. This study estimated T1w/T2w ratios in 42 left TLE (LTLE) and 42 right TLE (RTLE) patients and 41 healthy controls (HC). A T1w/T2w structural covariance network (SCN) was built by calculating correlations between any two regions across subjects and analysed by graph theory. Voxel-wise comparisons of T1w/T2w laterality were performed among the three groups. Compared to HC, both patient groups showed decreased T1w/T2w in frontotemporal regions, amygdala and thalamus; however, the LTLE showed lower T1w/T2w in left medial temporal regions than RTLE. Moreover, the LTLE exhibited decreased global efficiency compared with HC and more increased connections than RTLE. The laterality in putamen was differently altered between the two patient groups: higher laterality at posterior putamen in LTLE and higher laterality at anterior putamen in RTLE. This study demonstrated T1w/T2w reductions in frontotemporal and subcortical regions and extensive disconnections of SCN, providing evidence that TLE is a system disorder with widespread disruptions at regional and network levels. The putamen may play a transfer station role in damage spreading induced by epileptic seizures from the hippocampus.

## I. INTRODUCTION

Temporal lobe epilepsy (TLE) is the most common type of intractable epilepsy in adults [1]. Although magnetic resonance imaging (MRI) morphometric measures, such as cortical thickness or grey-matter volume, have demonstrated medial temporal atrophy, cortical thinning and subcortical abnormalities [2], a few studies found brain myelination alterations in TLE. For example, an earlier histopathological study of surgical specimens reported anomalies of cortical myelinated fibres in TLE [3]. A recent study has demonstrated atypical morphology of myelinated axons in TLE patients with temporo-polar blurring, using a combination of ex-vivo MRI and histological analysis [4]. To date, the non-invasive study of myelin changes in the human brain remains a challenge. Recently, a method based on the ratio of T1-weighted (T1w) and T2-weighted (T2w) magnetic resonance images provides insight in the characterization of microstructural myelination changes underlying brain cortical and subcortical grey-matter and white matter (WM) in vivo [5]. The T1w/T2w ratio, by simultaneously enhancing the sensitivity to myelin signal intensity while reducing the inter-subject signal intensity bias, can be used as a measure for detecting changes in myelination degree associated with diseases [5]. T1w/T2w ratio has been widely applied to study cortical and WM changes in ageing adults, multiple sclerosis and post-traumatic stress disorder [6, 7].

It is still unclear how these regions with myelination changes interact with each other. Do structural alterations across brain regions synchronously co-change in patients with TLE? Are the topological properties of the structural covariance network distinct between TLE and healthy controls? Although accumulated evidence has suggested different damages of grey-matter volume [8], cortical thickness [8], and the structural and functional network [9] between left TLE and right TLE, no studies have examined the impact of the seizure side on the myelination. As the most common drug-resistant type of epilepsy, TLE patients frequently receive an anterior temporal lobectomy (ATL) [1]. However, it remains unclear whether the myelination levels in the damaged regions before surgery improve after ATL.

This study applied the T1w/T2w analysis workflow to investigate in vivo the microstructural feature of brain tissue in a cohort of patients with left TLE, right TLE and healthy controls. Subsequently, we employed region of interest (ROI) analysis for the ROI-wise estimation of T1w/T2w in cortical grey-matter, subcortical grey-matter and WM. Simultaneously, to evaluate the synchronous changes of T1w/T2w across ROIs, we built a structural covariance network by calculating the correlations between any two ROIs across subjects in each group. We compared differences in the connections and topological properties of the structural covariance network among the left TLE, right TLE and healthy controls. In addition, we estimated the laterality map of T1w/T2w for homotopic voxels, which can achieve a voxel-wise comparison of T1w/T2w laterality among the left TLE, right TLE and healthy subjects. Finally, we investigated the longitudinal changes of T1w/T2w using another longitudinal sample including post-ATL TLE patients.

## II. Data

### A. Participants

This study included forty-two left TLE patients (LTLE group: 15 females, Age=25.29±7.96), 42 right TLE (RTLE group: 18 females, Age=26.96±8.33) and 41 healthy controls (HC group, 25 females, Age=29.37±10.63). All subjects were recruited from the West China Hospital and scanned by the MRI. All patients were diagnosed with unilateral TLE according to the International League Against Epilepsy (ILAE) [10]. A combination of ictal semiology, interictal and ictal EEG findings, MRI and PET/CT if applicable, was used to localize the seizure focus. All participants were right handedness. There was no significant difference in the subject age or gender among the three groups (P>0.05) (**Table 1**).

**Table 1.**
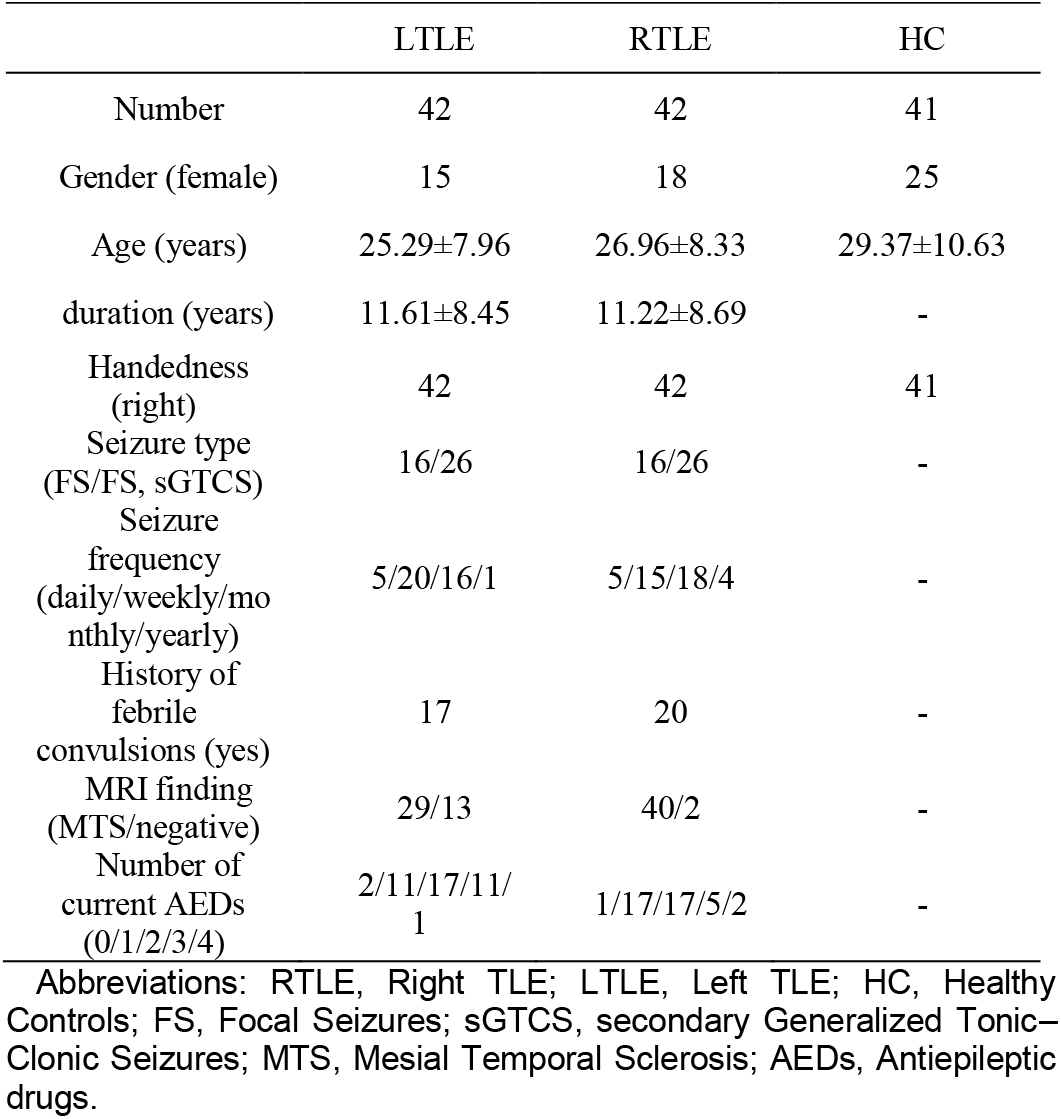
**Demographic and clinical variables**.

### B. Image acquisition

High-resolution T1-weighted images were acquired using spoiled gradient recalled sequence on a 3 T MRI system (Tim Trio; Siemens, Erlangen, Germany) with an eight-channel head coil at West China Hospital. The main parameters included: repetition time (TR) = 1900 ms; echo time (TE) = 2.26 ms; flip angle (FA) = 9°, field of view (FOV) = 256×256 mm ^2^; voxel size = 1.0×1.0×1.0 mm ^3^, 176 slices. T2-weighted images were acquired using a turbo spin echo (TSE) sequence (TR: 6100 ms; TE: 97 ms; FA: 120°; FOV: 230 mm; voxel size = 0.6×0.6×3.9 mm^3^; 35 slices). This study was approved by the local ethics committee and informed consent was obtained from all subjects. Ten of the 84 patients have been previously reported in our prior article which explored the dynamic changes of white matter microstructure measured by diffusion tensor imaging in mesial temporal lobe epilepsy patients following anterior temporal lobectomy [11]. Different than the previous research, the current study investigated abnormalities of myelin in temporal lobe epilepsy by using T1- and T2-weighted MRI data.

## III. Data Processing

A flowchart is provided in **Fig. 1**.

**Fig. 1.**
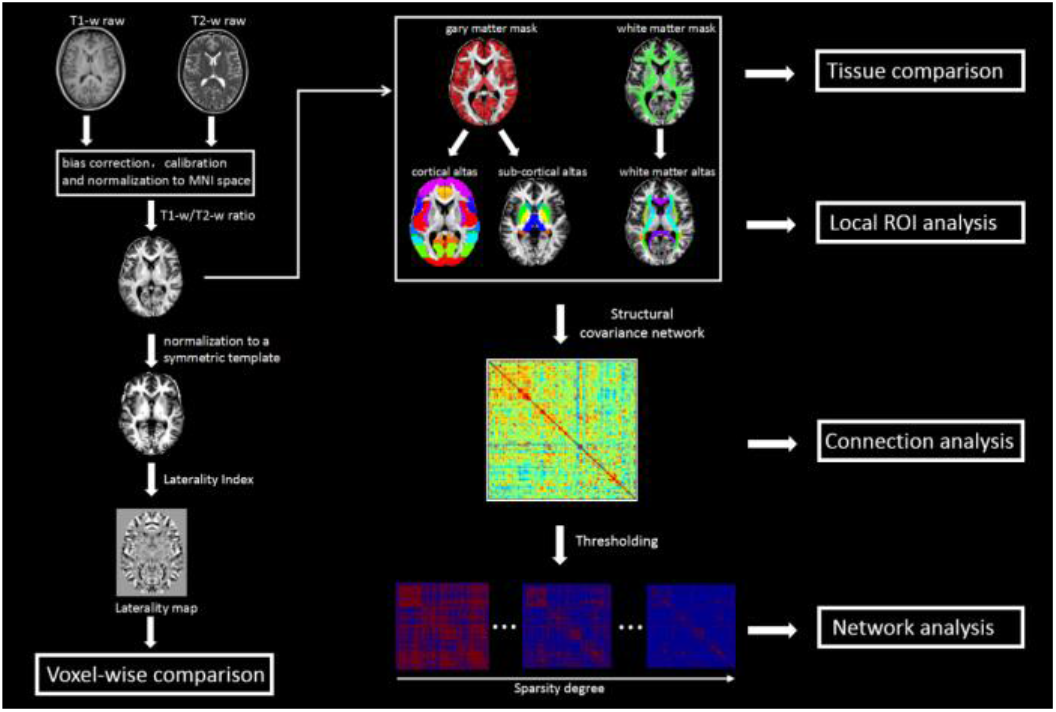
The flowchart of data processing. First, the T1w and T2w images were pre-processed by the bias correction, intensity calibration and normalization using the MR Tool-Multimodal Mapping (http://www.bindgroup.eu/wp-content/uploads/2017/02/mrtool-v1.3.1.zip) implemented in SPM12 (http://www.fil.ion.ucl.ac.uk/spm/software/spm12/) to obtain the T1w/T2w map. Second, the averaged T1w/T2w value was extracted from each ROI of JHU ICBM-DTI-81 white-matter atlas and the Harvard-Oxford grey-matter atlas. Third, the T1w/T2w structural covariance network (SCN) was built according to the strategy of the structural covariance network. The connection of the SCN was defined as the Pearson’s correlation coefficient across subjects between each pair of ROIs. Hence, the interregional correlation matrix (N×N, N=160) was obtained to represent the similarity or synchronized co-variations in T1w/T2w. Next, we used graph theory to characterize the SCN attributes. Finally, the laterality analysis was performed by first co-registering the T1w/T2w map into a symmetric template. Subsequently, we calculated the laterality index (LI) for each homotopic voxel and thus obtained a laterality map for each subject.

### A. T1w/T2w mapping

T1w and T2w images were pre-processed and combined, using a dedicated workflow outlined in previous studies [12]. This process included bias correction and intensity calibration on each of the T1w and T2 images and subsequent calculation of the ratio between the two images. In detail, each subject’s T2w image was co-registered to the T1w images using a rigid-body transformation. Second, bias correction was conducted on the T1w and T2w images separately. After correction for intensity non-uniformity, the T1w and T2w images were processed to standardize their intensity using a linear scaling procedure [12]. Subsequently, the ratio was calculated to obtain the T1w/T2w image. Finally, the T1w/T2w image was normalized to the Montreal Neurological Institute (MNI) space. The entire T1w/T2w image processing was performed using the MR Tool-Multimodal Mapping (Release 1.3.1, http://www.bindgroup.eu/wp-content/uploads/2017/02/mrtool-v1.3.1.zip), a MATLAB-based toolkit requiring SPM version 12 (http://www.fil.ion.ucl.ac.uk/spm/software/spm12/).

### B. Tissue and ROI analysis

The grey-matter (GM) and WM masks were defined based on the tissue probability map provided in SPM12 (threshold=50%). The average T1w/T2w intensity within GM and WM masks was calculated. To gain further insight to the precise regions, the WM component was refined into 48 ROIs (21 bundles in each hemisphere and 6 commissure bundles) using the JHU ICBM-DTI-81 WM atlas [13], and the GM cortical (96 ROIs) and subcortical (16 ROIs) components were refined based on the Harvard-Oxford GM atlas [14]. The average T1w/T2w intensity value was extracted from each ROI. ANCOVA was performed to examine the group difference among the LTLE, RTLE and HC, with age and gender as the covariates. The P value < 0.05/160 was considered significant for the multiple comparison correction (Bonferroni correction). In addition, the post hoc tests were performed for the comparisons between any two groups.

### C. T1w/T2w structural covariance network analysis

The T1w/T2w structural covariance network (SCN) was built as described in the previous study by Melie-Garcia [15]. Based on JHU ICBM-DTI-81 WM atlas and the Harvard-Oxford GM atlas, the entire brain was parcellated into 48 WM ROIs and 112 GM ROIs. The T1w/T2w data matrix was M × N, where M is the number of subjects and N represents the ROI number (N=160 in the current study). A linear regression was conducted on the T1w/T2w data matrix to remove the effects of gender, age and age^2^ [15]. The connection of the SCN was defined as the Pearson’s correlation coefficient across subjects between each pair of ROIs. Hence, the interregional correlation matrix (N×N, N=160) was obtained to represent the similarity or synchronized co-variations in T1w/T2w. To avoid the influence of the weak correlations, one sample t test was performed to determine the significant correlation (P<0.05, FWE correction). To compare the group differences in the connection strength, a permutation test was performed [16]. In brief, we randomly assigned the group labels across subjects and re-generated the SCN, which was repeated 100,000 times. We computed the difference of each connection between the two random groups and generated a distribution of the null hypothesis of equality in connection strength between groups. Based on the location of the real group difference of connection strength within the distribution of the null hypothesis, a p value was calculated for each connection.

Next, we used graph theory to characterize the SCN attributes. The sparse binary graphs were created using a range of sparsity degrees that varied from 0.1 to 0.4 with a step of 0.02 [15]. Subsequently, the network attributes, including clustering coefficient, characteristic path length, and global and local efficiency, were computed for the LTLE, RTLE and HC groups for each sparsity threshold. The formula and interpretations of graph measures can be found in prior research [17]. Finally, we calculated the area under the curve (AUC) for each network metric. The AUC metric can provide an integrated scalar for brain network topological characterization independent of a single threshold selection [18]. Statistical significance of the group differences was assessed using the permutation test, with a P-value < 0.05 [16]. In detail, group labels were randomly assigned across all subjects and the SCN attributes of each group was recalculated 10,000 times. The permutation processing allows for non-parametric estimation of the null distribution for the difference between any two groups.

### D. Laterality analysis

Due to the differences of geometric configuration between cerebral hemispheres, each subject’s T1w/T2w images were nonlinearly registered into a symmetrical template [19]. Referring to previous studies [19], we created the symmetrical template using the following steps. First, all subjects’ normalized T1w/T2w images were averaged to obtain a group mean template. Then, the mean group template was left-to-right flipped and re-averaged to create a symmetrical template [19]. In the symmetrical brain space, we calculated the laterality index (LI) for each homotopic voxel [20], that is:

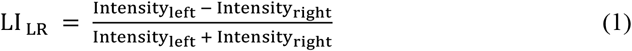

where Intensity_left_ represents the left T1w/T2w intensity and Intensity_right_ represents the right T1w/T2w intensity for each homotopic voxel.

By the LI calculation of each homotopic voxel, we obtained a laterality map for each subject. Voxel-wise ANCOVA was performed on the laterality maps to compare the differences among the LTLE, RTLE and HC, after controlling for the effects of gender and age. Multiple comparisons correction was performed using cluster-wise FDR correction (corrected P<0.05) [21].

## IV. Results

### A. Tissue and ROI analysis

Consistent with a previous study [22], tissue comparison confirmed that WM had higher T1w/T2w intensity than GM (**Fig. 2A**). In addition, we found T1w/T2w reductions in TLE for GM and WM, compared to controls (**Fig. 2A**).

**Fig. 2.**
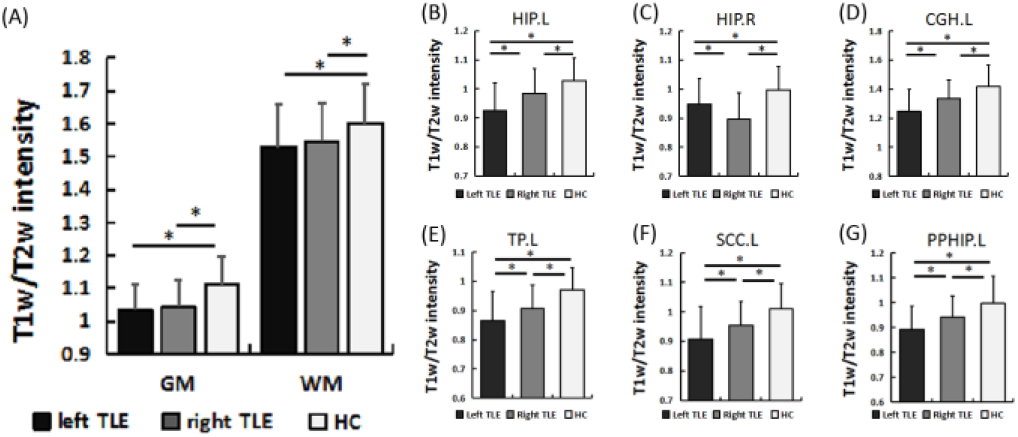
Differences of T1w/T2w intensity among the left TLE, right TLE and HC in (A) GM and WM masks, (B) left HIP, (C) right HIP, (D) left CGH, (E) left TP, (F) left SCC and (G) left PPHIP. The P<0.05/160 was considered significant for the multiple comparisons correction (Bonferroni correction). The error bars represent standard deviations. TLE, temporal lobe epilepsy; HC, healthy controls; GM, grey matter; WM, white matter; HIP, hippocampus; CGH, cingulum adjoining the hippocampus; TP, temporal pole; SCC, subcallosal cortex; PPHIP, posterior parahippocampal gyrus. * represents P<0.05, Bonferroni correction.

We then focused our investigation on specific brain regions within the WM and GM including sub-cortical and cortical regions. GM sub-cortical ROI comparisons showed that, compared with HC, both LTLE and RTLE groups had decreased T1w/T2w intensity in the left amygdala, left thalamus and bilateral hippocampus (**Fig. S1, Table S1**), with more severe reduction in the ipsilateral hippocampus (Fig. 2B and Fig. 2C) in each patient group. WM ROI analysis indicated lower T1w/T2w in bilateral cingulum adjoining the hippocampus (CGH) and fornix body (FIX) in both TLE groups (**Fig. S2, Table S2**). In left CGH, the LTLE exhibited reduced T1w/T2w intensity compared to the RTLE (**Fig. 2D**). Cortical ROI analysis found that compared with HC, both TLE groups had lower T1w/T2w in the temporal lobes (bilateral temporal pole [TP], left anterior superior temporal gyrus [ASTG], right posterior temporal fusiform cortex [PTFC], left posterior middle temporal gyrus [PMTG], left anterior inferior temporal gyrus [AITG], left posterior parahippocampal gyrus [PPHIP]), frontal lobes (left pars triangularis of inferior frontal gyrus [TriIFG], bilateral middle frontal gyrus [MFG], left subcallosal cortex [SCC], and left frontal pole [FP]), occipital lobes (left superior lateral occipital cortex [SLOC] and right occipital pole [OP]), parietal (left anterior supramarginal gyrus [ASMG]) and insular lobes (right insula [INS]) (**Fig. S3, Table S3**). Additionally, the LTLE showed more severe decreases in the left temporal pole (TP) (**Fig. 2E**), left SCC (**Fig. 2F**) and left PPHIP (**Fig. 2G**), compared with RTLE.

### B. SCN analysis

The edges of the SCN indicated significant correlations between T1w/T2w measures of specific ROIs across subjects. By comparing patients and healthy controls, the edges can be described as decreased connections (ROI-ROI correlation was significantly lower in patients compared with controls) and increased connections (ROI-ROI correlation was significantly higher in patients compared with controls) by the permutation test (**Fig. 3A**). In total, compared with HC, the LTLE group showed more changed connections than RTLE (59 vs. 35) (**Table 2**). Furthermore, there were more increased connections than decreased connections (29 vs. 19) in the LTLE group, while there were more decreased connections than increased connections (20 vs. 5) in the RTLE group (Table 2). In both LTLE and RTLE groups, there were more extensive changes in the cortical ROI to cortical ROI connection pattern, compared with other connection patterns (Table 2), such as subcortical ROI to subcortical ROI connection or WM ROI to WM ROI connection. In addition, there were more increased cortical-to-cortical connections in LTLE than RTLE (29 vs. 5), especially for the frontotemporal connectivity.

**Table 2.** **Decreased connections and increased connections of T1w/T2w structural covariance network in left TLE and right TLE**.

| Connection | Decrease |  | Increase |  |
| --- | --- | --- | --- | --- |
|  | LTLE | RTLE | LTLE | RTLE |
| Cortical ROI to cortical ROI connection | 19 | 20 | 29 | 5 |
| Subcortical ROI to Subcortical ROI connection | 1 | 0 | 3 | 2 |
| WM ROI to WM ROI connection | 2 | 3 | 0 | 2 |
| Cortical ROI to subcortical ROI connection | 2 | 1 | 0 | 2 |
| Cortical ROI to WM ROI connection | 3 | 0 | 0 | 0 |
| Subcortical ROI to WM ROI connection | 0 | 0 | 2 | 0 |
| Total | 25 | 24 | 34 | 11 |

**Fig. 3.**
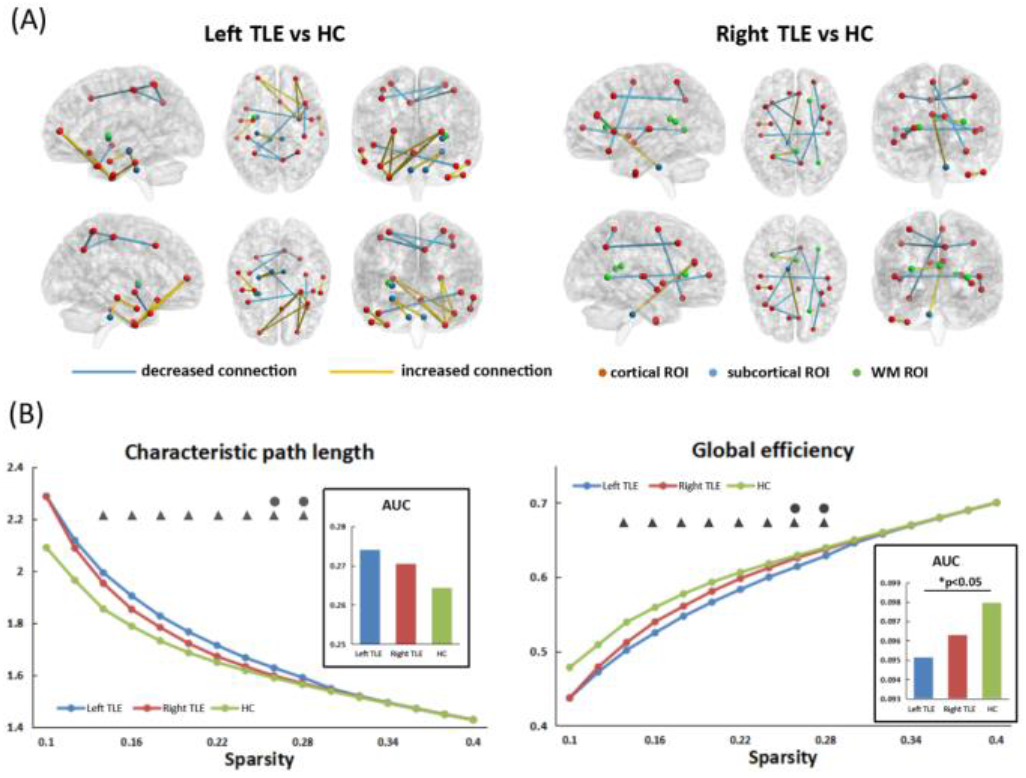
Changes of the T1w/T2w structural covariance network in left TLE and right TLE. (A) By comparing patients and healthy controls, the edges can be described as decreased connections (ROI-ROI correlation was significantly lower in patients compared with controls) and increased connections (ROI-ROI correlation was significantly higher in patients compared with controls). (B) Differences of attributes in the structural covariance network among the left TLE, right TLE and HC. The triangles represent significant differences between left TLE and HC (P<0.05). The circles represent significant differences between left TLE and right TLE (P<0.05). TLE, temporal lobe epilepsy; HC, healthy controls.

The permutation test revealed that compared with HC, only the LTLE group had higher characteristic path length and lower global efficiency for a range of sparsity threshold from 0.14 to 0.28 (All Ps<0.05) (**Fig. 3B**). Moreover, the comparisons between two patient groups showed higher characteristic path length and lower global efficiency in LTLE at the sparsity of 0.26 and 0.28 (P<0.05) (**Fig. 3B**). In addition, the AUC analysis showed that only the LTLE group exhibited a significant decrease (P<0.05) in the AUC of global efficiency and a marginally significant increase (P=0.050) in the AUC of characteristic path length, compared with HC (**Fig. 3B**). There was no significant difference among the healthy controls, LTLE and RTLE group, in terms of clustering coefficient and local efficiency (All Ps > 0.05).

### C. Laterality analysis

The ANCOVA on the LI_LR_ maps showed significant differences among the LTLE, RTLE and HC groups in anterior and posterior putamen, hippocampus and anterior temporal area (P<0.05, FDR correction) (**Fig. 4A**). The post hoc tests revealed that there was decreased LI_LR_ in LTLE and increased LI_LR_ in RTLE compared with HC (Ps<0.05, Bonferroni correction) (**Fig. 4B**). In detail, one-sample t-tests showed that the laterality of the anterior temporal area was significantly different from zero in all three groups (corrected Ps<0.05). The laterality of the hippocampus and posterior putamen existed in LTLE and RTLE (corrected Ps<0.05) but not in the HC. The laterality of the anterior putamen existed only in the RMTL (corrected P<0.05). Due to the LI_LR_ calculation (left-right), both LTLE and RTLE exhibited a consistent decrease of T1w/T2w LI_LR_ at the ipsilateral hippocampus. Except for the hippocampus and anterior temporal area, both TLE groups showed a T1w/T2w LI_LR_ decrease at the contralateral putamen (all Ps<0.05) (**Fig. 4B**).

**Fig. 4.**
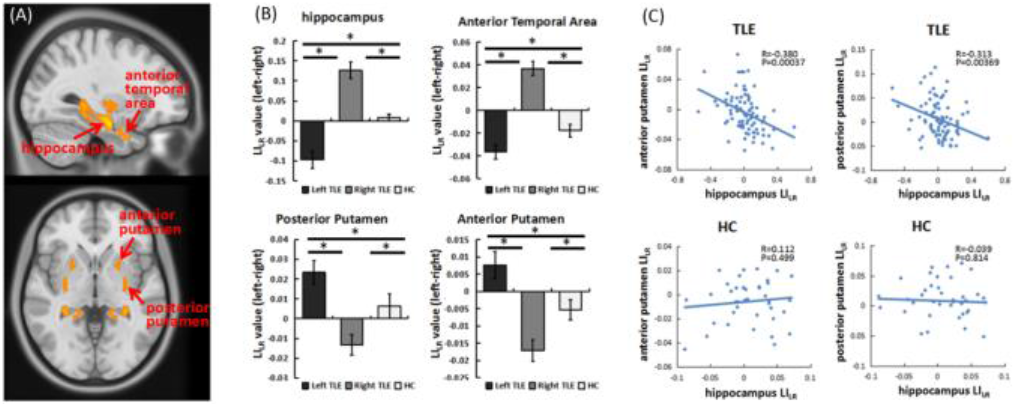
Results of laterality analysis in TLE. (A) Brain regions with significant differences of LI_LR_ maps among the left TLE, right TLE and HC groups by ANCOVA (P<0.05, FDR correction). (B) Altered LI_LR_ in the hippocampus and putamen in left TLE and right TLE compared with HC by post hoc analysis. (C) A significant correlation between hippocampal LI_LR_ and anterior and posterior putamen’s LI_LR_ in a cohort of all TLE patients but not in HC. TLE, temporal lobe epilepsy; HC, healthy controls; LI, laterality index. * represents P<0.05, Bonferroni correction.

As there are structural connections between the hippocampus and putamen, we further investigated whether a damaging synchronization between the hippocampus and putamen exists. We found a significant Pearson’s correlation between hippocampal LI_LR_ and anterior and posterior putamen LI_LR_ in a cohort of all TLE patients (anterior putamen: R=-0.380, P<0.001; anterior putamen: R=-0.313, P=0.004) but not in the HC (anterior putamen: R=0.112, P=0.499; anterior putamen: R=-0.039, P=0.814) (**Fig. 4C**).

In addition, we observed different alterations between putamen anterior and posterior regions in LTLE and RTLE. To better clarify the laterality changes in the putamen and perform a directed comparison between LTLE and RTLE, we redefined the laterality measure as the following equation:

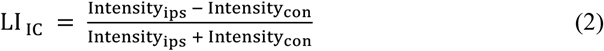

where Intensity_ips_ represents the ipsilateral T1w/T2w intensity and Intensity_con_ represents the contralateral T1w/T2w intensity for anterior and posterior putamen. A high value of the LI_IC_ represented a high T1w/T2w intensity on the contralateral putamen. We identified a significant interaction effect (F=4.26, P<0.05) between the side of epileptic focus (left or right) and the LI_IC_ of the putamen anterior or posterior (**Fig. S4**) using the repeated measured ANOVA. Specifically, the LTLE showed a higher laterality at the posterior putamen, while the RTLE exhibited a higher laterality at the anterior putamen (**Fig. S4**).

To further investigate the ATL influence on the putamen, we focused on the following questions: Are there longitudinal changes in the LI_IC_ of putamen after ATL? If so, how does the T1w/T2w of bilateral putamen change? We measured the longitudinal changes in another longitudinal cohort of 14 refractory TLE patients who have both pre-ATL and post-ATL MRI data (see Supplementary **Table S4** for details). Paired t-tests indicated increased LI_IC_ in both anterior (T=3.20, P=0.007) and posterior putamen (T=8.47, P<0.001) in the TLE patients after ATL (**Fig. S5**). Subsequently, to further determine which putamen subregions T1w/T2w changes contribute to the increased LI_IC_, we compared the longitudinal T1w/T2w intensity alterations between pre-ATL and post-ATL in ipsilateral and contralateral putamen subregions. We found increased T1w/T2w intensity only in the ipsilateral putamen after ATL (ipsilateral anterior putamen: T=2.91, P=0.012; ipsilateral posterior putamen: T=5.92, P<0.001) (**Fig. S6**).

## V. Discussion

This study used the T1w/T2w ratio to measure in vivo the brain microstructural changes in a large cohort of TLE patients, which allowed separate baseline analysis of LTLE and RTLE. Another cohort of TLE patients undergoing ATL surgery enables a further validation study of longitudinal changes induced by the ATL. First, compared to the HC, both LTLE and RTLE groups showed decreased T1w/T2w in cortical areas (mainly in frontotemporal regions), subcortical regions (hippocampus, amygdala and thalamus) and WM (CGH and FIX). Moreover, LTLE presented with lower T1w/T2w in left medial temporal region, compared to RTLE. Second, the SCN analysis indicated that compared to HC, both LTLE and RTLE groups had extensive changes of cortex-cortex connections. However, LTLE presented with more increased connections than RTLE. In addition, only the LTLE group exhibited a significant decrease in the global efficiency of SCN compared with HC. Third, the two TLE groups showed altered laterality of T1w/T2w in the hippocampus and putamen. Moreover, the T1w/T2w at the two regions exhibited a negative correlation in the patient cohort. Additionally, we found that the LTLE showed a higher laterality at the posterior putamen, where the RTLE exhibited a higher laterality at the anterior putamen. Lastly, we found increased T1w/T2w only in the ipsilateral putamen following the ATL.

### A. T1w/T2w intensity decreases in cortical, subcortical regions and WM in TLE

Across the two groups of TLE and HC, the comparisons using WM and GM as ROIs suggested a global reduction of T1w/T2w intensity in TLE. This result is consistent with current studies suggesting that TLE is a system-level disorder, with both GM and WM disruptions as the foundation for the clinical outcome of the pathology [23]. Regarding structural impairments in GM cortices, we found prominent alterations in TLE patients to be localized in the frontal and temporal lobes, which were consistent with previous studies revealing that abnormalities of frontal-temporal networks may be linked to cognitive impairments and pathological outcomes in TLE [24]. Regarding subcortical structures, we found T1w/T2w decreases in the hippocampus, amygdala and thalamus. The results were consistent with cumulative evidence indicating a key role of a thalamotemporal network in the pathologic alterations of TLE [25]. Regarding the WM results, the current study identified a reduction of T1w/T2w intensity in the WM tracts of CGH and FIX in the TLE patients. Several studies have also reported consistent WM aberrations in hippocampal WM pathways [26]. Previous studies using diffusion tensor imaging have observed widespread microstructural organizational changes, such as reduced fractional anisotropy (FA), which is possibly linked to myelination alterations [27]. Previous work has reported aberrant myelin content in children with epilepsy [28]. Moreover, a recent review also suggested a relationship between pathophysiology of epilepsy and myelin abnormalities [29]. The present findings provided brain microstructural changes of T1w/T2w ratios in TLE, using a combination of two MRI measures [12]. The observed changes in T1w/T2w used in this study could be due to changes in myelination of TLE patients, but it should be recognized that other microstructural changes may be responsible.

### B. T1w/T2w structural covariance network is distinct in left and right TLE

The structural covariance network method has been widely used to investigate the concurrent changes of brain structures (i.e., cortical thickness) in normal and pathological brain [30]. To characterize the concurrent changes in T1w/T2w across brain anatomical structures in vivo, this study built a SCN by calculating the synchronicity of the variations in T1w/T2w between anatomical regions. The T1w/T2w covariance network analysis pointed to more severe and more extensive changes in LTLE than RTLE. Such distinct network alterations agree with several structural studies of GM and WM [31, 32]. For example, morphological analysis on GM volume showed a stronger reduction in patients with left-sided seizure origin [33]. Investigations of FA in WM tracts identified more widespread deficits in LTLE [34]. Focke et al. also found that the WM tract of arcuate fasciculus was more affected in LTLE [35]. Wang and colleagues reported aberrant topological patterns of metabolic covariance networks in TLE [36]. In addition, Campos et al. indicated that LTLE had a more intricate bilateral bi-hemispheric dysfunction compared to RTLE using resting-state fMRI [9]. In total, our findings identified a distinct pathological network of T1w/T2w, which exhibited more connectome alterations in LTLE than RTLE, supporting previous suggestions that LTLE is a more severe network disease than RTLE [31].

The topological properties of the structural covariance network were also altered in TLE. The “characteristic path length” was statistically higher in LTLE. The increased “characteristic path length” was accompanied by a decreased “global efficiency” in principle. According to the graphical theory, higher “characteristic path length” represents higher “wiring cost” for information communication within the network. Additionally, human brain studies also demonstrated that the interpretation is reasonable. For example, Melie-Garcia and colleagues reported an increasing characteristic path length in an old-age group compared to young-age subjects, suggesting that worse network efficiency is related to poor performances in older subjects [15]. The altered topological organization in TLE patients indicated worse network efficiency, suggesting insufficient information interactions in the brain. These findings provided further support for the hypothesis that abnormalities of brain network efficiency in LTLE may be more serious than in RTLE [37].

### C. Laterality of T1w/T2w analysis: the putamen may play a crucial role in controlling epileptic seizures

Asymmetry is used for pathological detection for several reasons. First, the degree of asymmetry is commonly reviewed by expert radiologists who visually inspect the neuroimaging data in search of pathology. Second, asymmetry measurement can reduce confounding factors from differences in diseases, scanner parameters and neuroimaging modalities, by using each subject as a control for itself. Third, previous studies have shown that asymmetric measures are more sensitive to pathological characterization than raw voxel values [38].

In TLE, previous studies have shown asymmetric atrophy in mesial temporal structures and asymmetric reduction of glucose metabolism in the temporal lobes [38]. A recent study found that TLE patients showed greater grey matter volume in contralateral hippocampus and subfields, indicating volumetric asymmetry in TLE [39]. Resting-state fMRI studies also reported decreased ipsilateral functional connectivity within the mesial temporal lobes in TLE [40]. Consistent with these findings, we observed asymmetric anomalies related to pathological changes in the hippocampus, using the ratio of T1w/T2w images. In addition, the current study found asymmetric alteration of T1w/T2w in the putamen in TLE. The putamen, as a subcortical region, is a principal anatomical component of the basal ganglia, which receives inputs from the cortical areas and sends information back to the cortex via the thalamus [41]. The basal ganglia shows intimate structural connections with the brain limbic system, especially the hippocampus [42]. Reduction of grey-matter volume has also been reported in the basal ganglia, including the putamen in TLE patients [43]. Additionally, aberrant functional connectivity in the basal ganglia and hippocampus has been noted previously in TLE [44]. Furthermore, we found that the T1w/T2w values of the putamen negatively correlated with that of the hippocampus. According to the structural connection and functional association between the hippocampus and putamen, we hypothesized that the putamen may play a transfer station role in damage spreading induced by epileptic seizures from the hippocampus. The hypothesis was further supported by the finding that the T1w/T2w of the putamen increased after the ATL. These results are consistent with previous studies reporting that the basal ganglia plays an important role in control of the epileptic activity through the brain [45]. However, we found the myelination improvement only appeared in the ipsilateral putamen, which may suggest a lateralization effect of the putamen improvement induced by the ATL. These findings may guide the development of new intervention targets or treatment strategies that may modulate morphological reorganization in specific subcortical nuclei.

In addition, we found different patterns of T1w/T2w laterality in putamen subregions between LTLE and RTLE, revealed by a higher laterality at the posterior putamen in the LTLE, but a higher laterality at the anterior putamen in the RTLE. This finding provided evidence for identifiable differences of myelination damages in the putamen between LTLE and RTLE.

### D. Limitations

The current study had several limitations. First, the present study used the T1w/T2w ratio method to characterize the brain changes of myelination. It is necessary to investigate the reproducibility using other measures of myelination mapping, such as magnetization transfer ratio (MTR), although previous studies have demonstrated a high degree of correlation between MTR and T1w/T2w ratio [46]. Second, myelination largely contributes to the ratio of the T1w/T2w images; however, some factors, including inflammation, oedema, metabolism, atrophy or iron accumulation, may also contribute to the T1w/T2w signals [12]. The T1w/T2w ratio used in the current study is an estimate of T1w/T2w, not a fully quantitative measure of it (i.e., quantitative T1 and T2 relaxometry was not performed). Third, the cohorts of patients before and after ATL surgery enable a baseline comparison and a further investigation of longitudinal changes following surgery. Not all patients underwent the ATL surgery. In addition, the dynamic brain changes should be further investigated on data from multiple postoperative time points. Finally, disease duration and medication use may also affect the secondary changes in the T1/T2 ratio map. A larger sample size with greater statistical power is needed to further verify the significance of the group differences.

## VI. Conclusion

This study demonstrated T1w/T2w reductions in the frontotemporal and thalamic regions and extensive disconnections of a T1w/T2w structural covariance network, providing evidence that TLE is a system disorder with widespread disruptions at regional and network levels. The T1w/T2w laterality of the putamen at baseline was altered and negatively related with the hippocampus in TLE. Furthermore, ATL surgery increased T1w/T2w values in the ipsilateral putamen. This finding suggested that the putamen may play a transfer station role in damage spreading induced by epileptic seizures from the hippocampus. Last, we found identifiable differences of T1w/T2w changes between LTLE and RTLE, revealed by more increased connections and decreased global efficiency of a covariance network in LTLE.

## Data Availability

The data supporting the findings of this study are available from the corresponding author upon reasonable request.

