## Supplementary Materials for "In vivo Characterization of MRI-based T1w/T2w Ratios and Covariance Network in Temporal Lobe Epilepsy"

<u>**Supplementary material for**</u>

**Running title:** *Changes of T1w/T2w in temporal lobe epilepsy*

**Keyword:** *Myelin; Temporal lobe epilepsy; Magnetic resonance image; Structural covariance; Putamen; Structural connectivity*

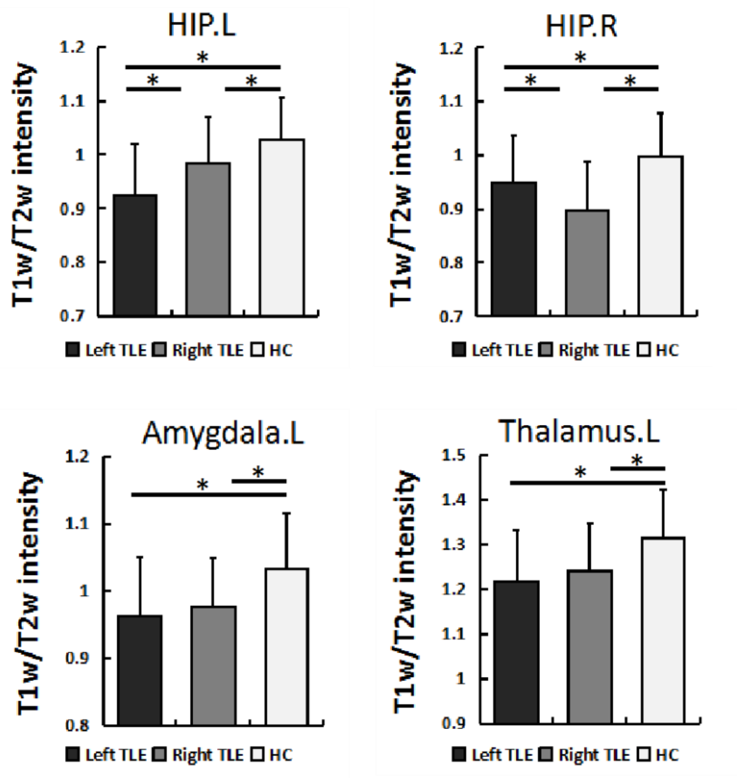


**Figure S1.** Differences of T1w/T2w intensity among the left TLE, right TLE and healthy controls (HC) in the grey matter sub-cortical regions of left amygdala, left thalamus and bilateral hippocampus

**Table S1.** Regions with significant differences of T1w/T2w intensity among the left TLE, right TLE and healthy controls in the grey matter sub-cortical regions

| Regions | Mean value | | | ANCOVA | | Post hoc: p-value | | |
|---|---|---|---|---|---|---|---|---|
| | LTLE | RTLE | HC | F | P | LTLE vs RTLE | LTLE vs HC | RTLE vs HC |
| Left HIP | 0.935 | 0.985 | 1.028 | 14.6 | <0.00001 | 0.0019 | <0.00001 | 0.028 |
| Right HIP | 0.948 | 0.897 | 0.997 | 13.7 | <0.00001 | 0.0088 | 0.011 | <0.00001 |
| Left amygdala | 0.962 | 0.976 | 1.033 | 8.99 | 0.00023 | 0.422 | 0.00010 | 0.0017 |
| Right Thalamus | 1.216 | 1.240 | 1.314 | 8.82 | 0.00026 | 0.318 | 0.00009 | 0.0029 |

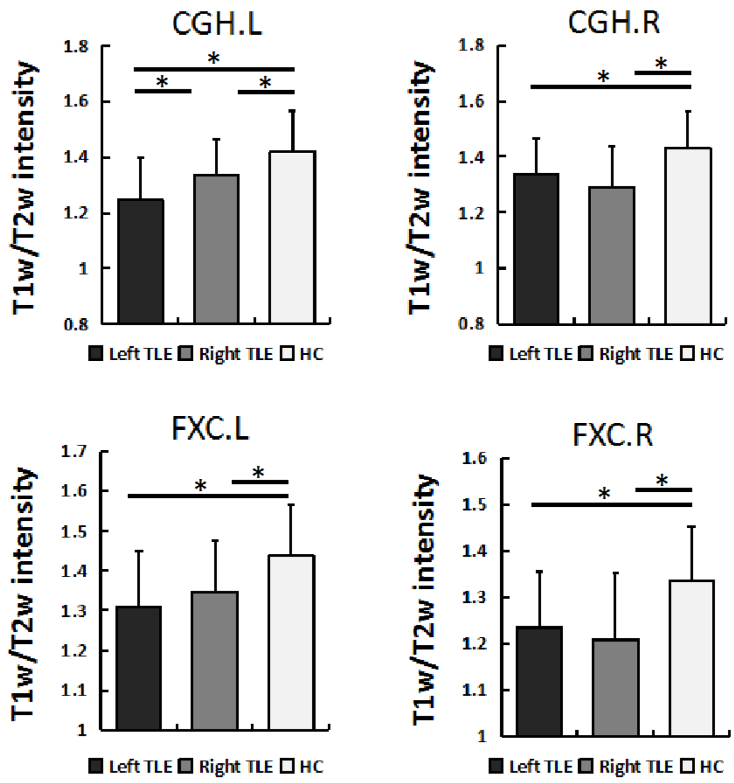


**Figure S2.** Differences of T1w/T2w intensity among the left TLE, right TLE and healthy controls (HC) in the white matter regions of bilateral cingulum adjoining the hippocampus (CGH) and fornix body (FIX)

**Table S2.** Regions with significant differences of T1w/T2w intensity among the left TLE, right TLE and healthy controls in the white matter regions

| Regions | Mean value | | | ANCOVA | | Post hoc: p-value | | |
|---|---|---|---|---|---|---|---|---|
| | LTLE | RTLE | HC | F | P | LTLE vs RTLE | LTLE vs HC | RTLE vs HC |
| Left CGH | 1.246 | 1.337 | 1.417 | 15.0 | <0.00001 | 0.004 | <0.00001 | 0.011 |
| Right CGH | 1.336 | 1.291 | 1.430 | 11.1 | 0.00004 | 0.131 | 0.002 | <0.00001 |
| Left FXC | 1.307 | 1.347 | 1.437 | 10.4 | 0.00007 | 0.178 | 0.00002 | 0.002 |
| Right FXC | 1.235 | 1.209 | 1.334 | 11.1 | 0.00004 | 0.339 | 0.0006 | 0.00002 |

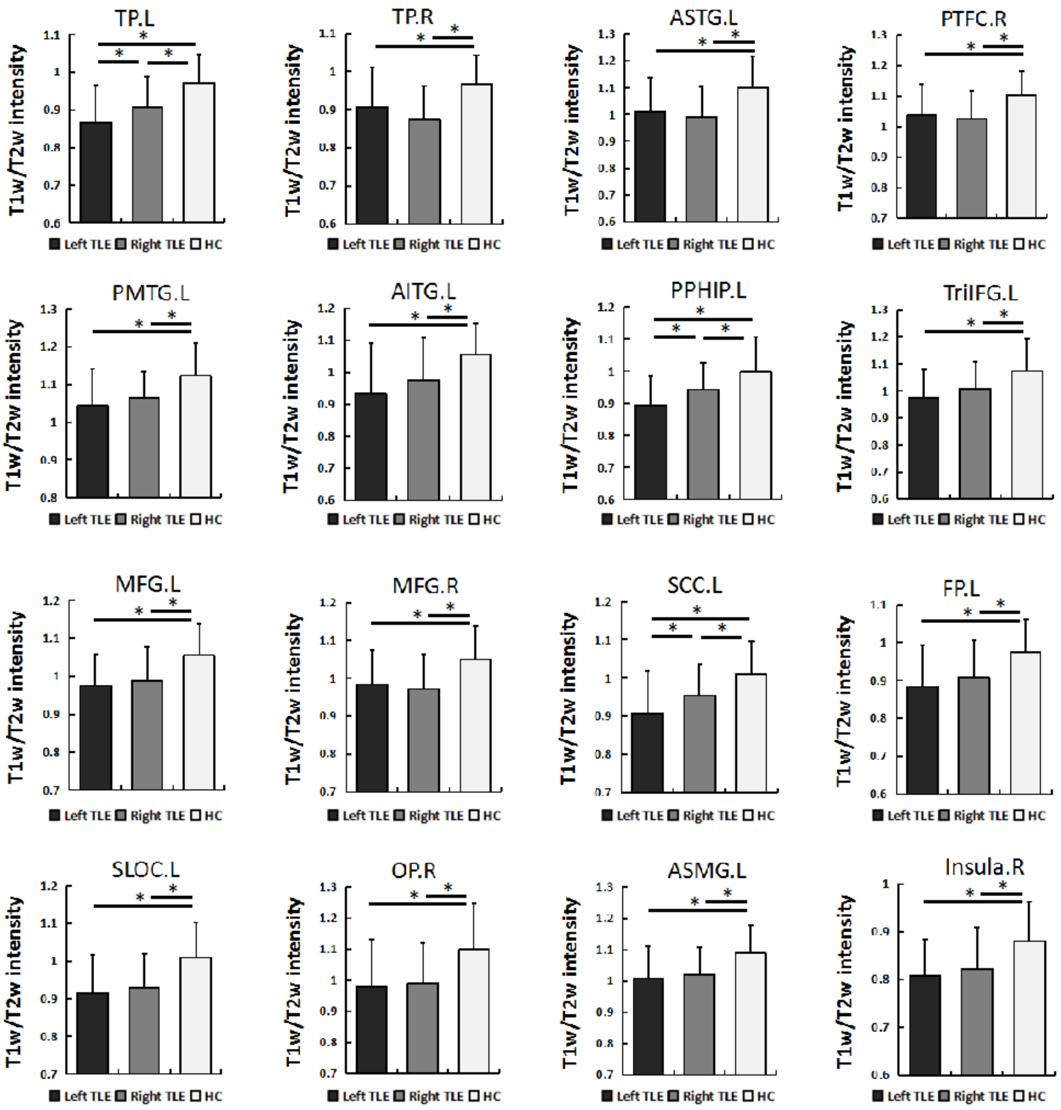


**Figure S3.** Differences of T1w/T2w intensity among the left TLE, right TLE and healthy controls (HC) in the cortical regions of temporal lobes (bilateral temporal pole [TP], left anterior superior temporal gyrus [ASTG], right posterior temporal fusiform cortex [PTFC], left posterior middle temporal gyrus [PMTG], left anterior inferior temporal gyrus [AITG], left posterior parahippocampal gyrus [PPHIP]), frontal lobes (left pars triangularis of inferior frontal gyrus [TriIFG], bilateral middle frontal gyrus [MFG], left subcallosal cortex [SCC], left frontal pole [FP]), occipital lobes (left superior lateral occipital cortex [SLOC] and right occipital pole [OP]), parietal (left anterior supramarginal gyrus [ASMG]) and insular lobes (right insula [INS])

**Table S3.** Regions with significant differences of T1w/T2w intensity among the left TLE, right TLE and healthy controls in the cortical regions

| Regions | Mean value | | | ANCOVA | | Post hoc: p-value | | |
|---|---|---|---|---|---|---|---|---|
| | LTLE | RTLE | HC | F | P | LTLE vs RTLE | LTLE vs HC | RTLE vs HC |
| Left TP | 0.865 | 0.906 | 0.972 | 16.3 | <0.00001 | 0.028 | <0.001 | 0.001 |
| Right TP | 0.906 | 0.873 | 0.967 | 11.4 | 0.00003 | 0.098 | 0.003 | <0.001 |
| Left ASTG | 1.013 | 0.991 | 1.102 | 10.3 | 0.00007 | 0.400 | 0.001 | <0.001 |
| Right PTFC | 1.035 | 1.025 | 1.103 | 8.78 | 0.00027 | 0.598 | 0.001 | <0.001 |
| Left PMTG | 1.042 | 1.064 | 1.123 | 9.81 | 0.00011 | 0.234 | <0.001 | 0.002 |
| Left AITG | 0.933 | 0.976 | 1.055 | 9.21 | 0.00019 | 0.139 | <0.001 | 0.007 |
| Left PPHIP | 0.892 | 0.943 | 0.996 | 11.8 | 0.00002 | 0.018 | <0.001 | 0.015 |
| Left TriIFG | 0.972 | 1.009 | 1.074 | 9.39 | 0.00016 | 0.121 | <0.001 | 0.007 |
| Left MFG | 0.976 | 0.988 | 1.055 | 10.4 | 0.00007 | 0.495 | <0.001 | 0.001 |
| Right MFG | 0.984 | 0.972 | 1.049 | 8.72 | 0.00029 | 0.543 | 0.001 | <0.001 |
| Left SCC | 0.908 | 0.953 | 1.011 | 12.9 | <0.00001 | 0.027 | <0.001 | 0.005 |
| Left FP | 0.882 | 0.907 | 0.975 | 9.62 | 0.00013 | 0.250 | <0.001 | 0.002 |
| Left SLOC | 0.914 | 0.927 | 1.011 | 12.2 | 0.00001 | 0.532 | <0.001 | <0.001 |
| Right OP | 0.979 | 0.990 | 1.099 | 8.79 | 0.00027 | 0.717 | <0.001 | 0.001 |
| Left ASMG | 1.009 | 1.020 | 1.090 | 9.06 | 0.00021 | 0.605 | <0.001 | 0.001 |
| Right Insula | 0.808 | 0.822 | 0.881 | 9.14 | 0.00020 | 0.464 | <0.001 | 0.001 |

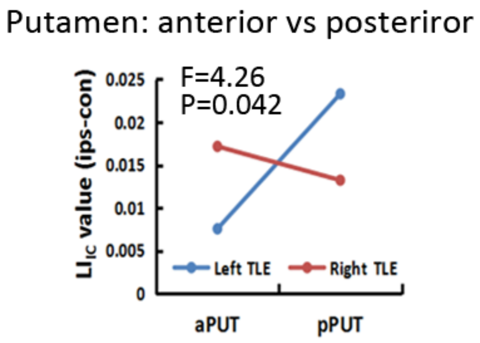


**Figure S4.** A significant interaction effect between the seizure side of TLE (left TLE or right TLE) and the $LI_{IC}$ of the putamen anterior or posterior by using the repeated measured ANOVA. The left TLE showed a higher laterality at the posterior putamen, while the right TLE exhibited a higher laterality at the anterior putamen.

### Longitudinal sample

Dataset-2 (longitudinal sample) included 14 TLE patients (10 left TLE, 6 females, Age=23.40±7.62; 4 right TLE, 2 females, Age=25.00±5.35) who underwent unilateral ATL based on comprehensive preoperative evaluations by epileptologists, neurosurgeons and radiologists. Patients were followed-up every three months after surgery and surgical outcome was evaluated based on the ILAE. MRI images were scanned at pre-surgically in all 14 patients and at three months after surgery in seven patients, six months in three, and 12 months in four.

**Table S4.** Characteristics of patients who received anterior temporal lobectomy resection (ATL) surgery.

| **Variable** | **ILAE Class I (n=14)** |
|---|---|
| Age (y) | 23.7 (15-41) |
| Sex | 8 female, 6 male |
| Age at onset (y) | 12.8 (1-28) |
| Epilepsy duration (y) | 10.9 (2-21) |
| Laterality | 10 left, 4 right |
| MTS on MRI scans | 12 MTS, 2 non-MTS |
| Handedness | Right, n=14 |

Note: MRI images were scanned at the baseline (pre-ATL) in all 14 patients and at the post-ATL three months in seven patients, six months in three, and 12 months in four.

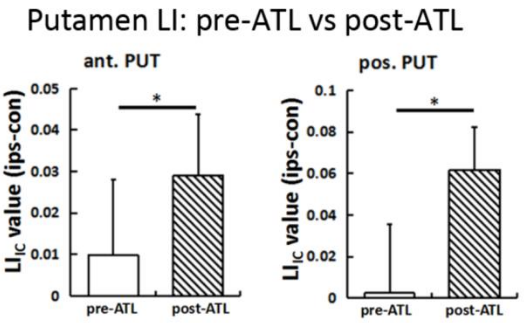


**Figure S5.** Increased $LI_{IC}$ in both anterior (T=3.20, P=0.007) and posterior putamen (T=8.47, P<0.001) in the TLE patients after ATL by paired t-test in a longitudinal cohort of 14 refractory TLE patients.

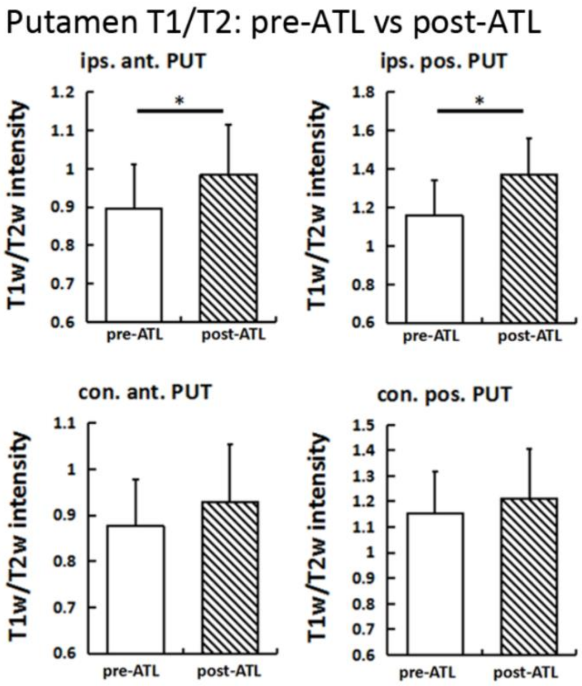


**Figure S6.** Increased T1w/T2w intensity between pre-ATL and post-ATL only in the ipsilateral putamen subregions in a longitudinal cohort of 14 refractory TLE patients.
